# Redefining Left Bundle Branch Block from High-density Electroanatomical Mapping

**DOI:** 10.1101/2023.02.20.23286204

**Authors:** Jun-Hua Zou, Bao-Tong Hua, Xiao-Xia Shao, Chao Wang, Hao Li, Ya-Nan Lu, Xin Tian, Zhi-Xuan Li, Li-Jin Pu, Jing Wang

## Abstract

**Background:** The current ECG criteria for diagnosing left bundle branch block (LBBB) still cannot fully differentiate between true and false blocks. The absence or presence of an LBBB is key in improving the response rate of clinical cardiac resynchronization therapy (CRT).

**Methods:** We hypothesized that the notch width of the QRS complex in the lateral leads (I, avL, V5, V6) on the LBBB-like ECG could further confirm the diagnosis of true complete left bundle branch block (t-LBBB). We performed high-density, three-dimensional electroanatomical mapping in the cardiac chambers of 37 patients scheduled to undergo CRT and whose preoperative electrocardiograms met the ACC/AHA/HRS guidelines for the diagnosis of complete LBBB. If the left bundle branch potential could be mapped from the bottom of the heart to the apex on the left ventricular septum, it was defined as a false complete left bundle branch block (f-LBBB). Otherwise, it was categorized as a t-LBBB. We compared the clinical characteristics, the real-time correspondence between the spread of ventricular electrical excitation and the QRS wave, the QRS notch width of the lateral leads (I, avL, V5, V6), and the notch width/left ventricular end-diastolic diameter (Nw/LVd) ratio between the two groups. Through ROC correlation analysis of Nw/LVd and t-LBBB, the sensitivity, specificity, and cut-off value of Nw/LVd diagnostic authenticity were obtained.

**Results:** Twenty-five patients were recruited to the t-LBBB group, and 12 to the f-LBBB group. In the t-LBBB group, the first peak of the QRS notch corresponded to the depolarization of the right ventricle and septum, the trough corresponded to the depolarization of the left ventricle across the left ventricle, and the second peak corresponded to the depolarization of the left ventricular free wall. In the f-LBBB group, the first peak corresponded to the depolarization of the right ventricle and most of the left ventricle, the second peak corresponded to the depolarization of the latest, locally-activated myocardium of the left ventricle, and the trough was caused by the off-peak delayed activation of the left ventricle. The QRS notch width (45.2 ± 12.3 ms vs. 52.5 ± 9.2 ms, *P*<0.05) and the Nw/LVd (0.65 ± 0.19 ms/mm vs. 0.81 ± 0.17 ms/mm, *P*<0.05) were compared between the two groups. Through ROC correlation analysis, the sensitivity (88%), specificity (58%), and cut-off value (0.56) for Nw/LVd diagnosis of t-LBBB was obtained.

**Conculuion:** Using the current diagnostic criteria of LBBB, increasing the Nw/LVd value can diagnose LBBB more effectively.

## Introduction

Left bundle branch block (LBBB) is closely associated with the progression and prognosis of chronic cardiovascular disease. It is an independent predictor of heart failure, sudden death, cardiovascular death, and all-cause mortality. Due to the change in depolarization direction, it induces interventricular desynchronization, which —especially in cases of true left bundle branch block (t-LBBB)—has a significant impact on hemodynamics (1–3). Cardiac resynchronization therapy (CRT) is currently an important way to treat heart failure presenting with LBBB. Left bundle branch pacing, lateral vein pacing, and His bundle pacing are used to achieve CRT. A key factor affecting the efficacy of CRT is the correct identification of t-LBBB. Unfortunately, according to the existing LBBB diagnostic criteria, the non-response rate to CRT is still as high as 30% (4).

In t-LBBB, the excitation of the left ventricle relies on the transseptal conduction of right ventricular electrical activity. Right ventricular electrical activity simultaneously spreads to the right ventricular free wall and septum, to the earliest breakthrough point of the left endocardium. This process takes around 40–50 ms. Immediately afterward, the electrical activity spreads from the septum to the heart chamber, which takes around 90–100 ms, so patients with LBBB have a longer QRS duration (5). On this basis, the diagnostic criteria for LBBB are constantly being improved. Currently, the most used LBBB diagnostic standard (ACC/AHA/HRS or Strauss criteria) has better sensitivity than the original diagnostic criteria, but its specificity is still too low to accurately diagnose t-LBBB (6). Approximately one third of patients diagnosed with LBBB by conventional ECG criteria are likely to receive a false positive diagnosis (7). Therefore, the accuracy of LBBB diagnosis must be further improved.

## Methods

### Study population

This was a prospective study of 37 surgical patients with heart failure (HF), reduced left ventricular ejection fraction (LVEF), and ECG suggestive of LBBB with indications for CRT. The three preoperative surface electrocardiograms of the patients met the ACC/AHA/HRS guidelines for LBBB diagnostic criteria (QRS duration ≥120 ms; notched or slurred R-wave in leads I, aVL, V5, and V6; absent Q-wave in I, V5, and V6; R peak time >60 ms in V5 and V6; ST and T-wave usually opposite to QRS). All participants gave written informed consent. Patients whose preoperative body surface electrocardiogram did not meet the diagnostic criteria of Strauss LBBB, patients with right bundle branch block, patients who refused to undergo CRT surgery, and patients with other organic heart diseases were excluded.

Approval for the study was gained from the Ethical Review Board of the First Affiliated Hospital of Medical University. We ensured that our research complied with the relevant legal and ethical regulations. We obtained informed consent from everyone meeting the inclusion criteria, and they signed the relevant documents.

### Study Approval

Approval for the study was gained from the Ethical Review Board of the First Affiliated Hospital of Medical University. We ensured that our research complied with the relevant legal and ethical regulations. We obtained informed consent from everyone meeting the inclusion criteria, and they signed the relevant documents.

### Three-dimensional electroanatomical maps of the LV

We performed femoral artery puncture and sheath placement on patients who met the inclusion criteria and were scheduled for CRT implantation by the method described (8), then sent in the PentaRay multi-electrode mapping catheter (PentaRay NAV eco high-density mapping catheter, Biosense Webstar, Irvine, CA, USA) or Orion basket mapping catheter (Constellation, Boston Scientific, Natick, MA, USA). We mapped left ventricular activation with the corresponding support system: a non-fluoroscopic navigation system (Fast Anatomical Mapping, CARTO 3®, version 6, Biosense Webster Inc., CA, USA) or electromagnetic navigation system (Rhythmia, Boston Scientific, Washington, DC, USA).

Through high-density three-dimensional electroanatomical mapping, if the left bundle branch potential could be mapped from the bottom of the heart to the apex of the left ventricular septum, it was defined as a false complete left bundle branch block (f-LBBB). If the left bundle branch region did not show significant potentials, the left bundle branch potential lagged behind the QRS complex in mapping, and mechanical stimulation of this region occasionally advanced the left bundle branch potential and induced subsequent premature ventricular contractions. At the same time, electrical stimulation was performed in the middle and upper regions of the left bundle branch. If the surface lead V1 was in the form of QR or rSR, it suggested the existence of t-LBBB, which was defined as the t-LBBB group.

In addition, we used high-density three-dimensional electrical mapping to describe the electrical activation of the left ventricle, compared the relationship between the spread of endocardial electrical activation and the notch of the body surface ECG in real time, and analyzed the mechanism of notch generation in the two groups.

### Surface ECG analysis

Patients scheduled to undergo CRT implantation, and whose preoperative ECG conforms to the 2009 ACC/AHA/HRS guidelines for the diagnosis of LBBB, should undergo no fewer than three ECG examinations and no fewer than two cardiac color Doppler ultrasound examinations. The ECG was analyzed simultaneously by two independent observers. We selected 10 adjacent QRS waves in the lateral wall leads (I, avL, V5, V6) of the preoperative ECG for measurement. The QRS duration was obtained by measuring the distance from the QRS origin to the J point and taking the average value. In the absence of premature beats or other arrhythmias, we calculated the timing of the two peaks in the QRS wave of the lateral leads (I, avL, V5, V6) and took the average to obtain the notch width (Nw). The ECG results measured by two observers were compared. The difference between the two measurements of the left ventricular end-diastolic diameter should be less than 2 mm, and the average value was taken to obtain the ratio of the notch width to the left ventricular end-diastolic diameter (Nw/Lvd).

### Statistical analysis

The measurement data conforming to normal distribution is represented by the mean ± standard deviation; the comparison between two groups is represented by the t-test; the statistics are represented by t. The enumeration data are represented by the frequency (rate/component ratio), and the comparison between the groups is represented by the χ^2^ test or Fisher’s exact test. We corrected the possible confounding factors by multivariate logistic regression analysis and adjusted the odds ratio (OR) and 95% confidence interval (CI) of the t-LBBB group by increasing the standard deviation of Nw/Lvd. We calculated the receiver operating characteristic (ROC) curve and the area under the ROC curve (AUC) to evaluate the predicted value. All data were statistically analyzed using SPSS 26.0.

## Results

### Patients

A total of 37 patients were enrolled in this study, all of whom underwent high-density three-dimensional electroanatomical mapping. Among them, 25 patients were confirmed as t-LBBB patients, and 12 were confirmed as f-LBBB patients. The baseline characteristics of the patients are summarized in **Table 1**. In the t-LBBB group, 18 people were diagnosed with dilated cardiomyopathy, and four were diagnosed with ischemic cardiomyopathy. The average preoperative QRS width was 160.5 ± 15.3 ms, and the average QRS notch width was 52.5 ± 9.2 ms. In the f-LBBB group, 10 cases were diagnosed as dilated cardiomyopathy, and two were diagnosed as alcoholic cardiomyopathy. The average preoperative QRS width was 175.6 ± 24.7 ms, and the average notch width was 45.2 ± 12.3 ms. Baseline age, sex, echocardiographic measurements, and other factors did not differ significantly between the t-LBBB and f-LBBB groups.

**Table 1:**
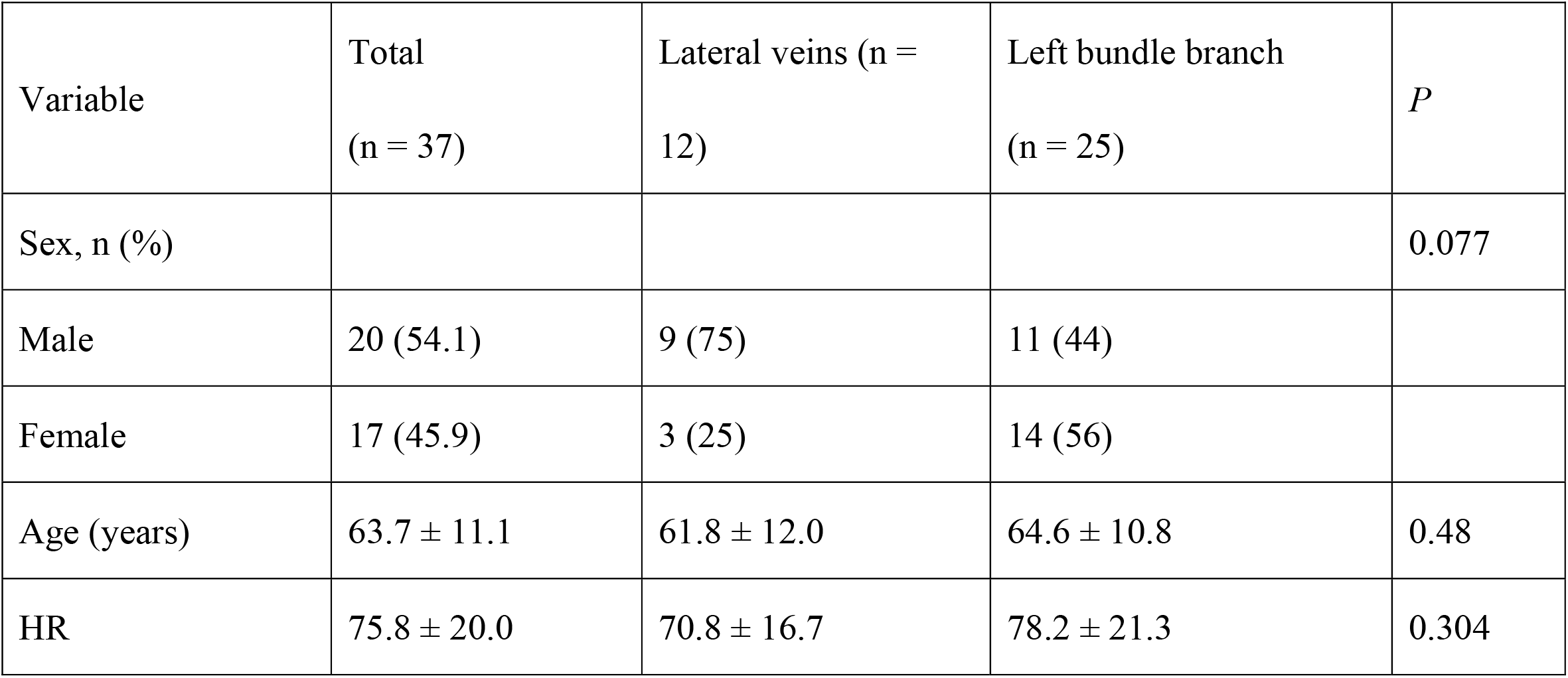
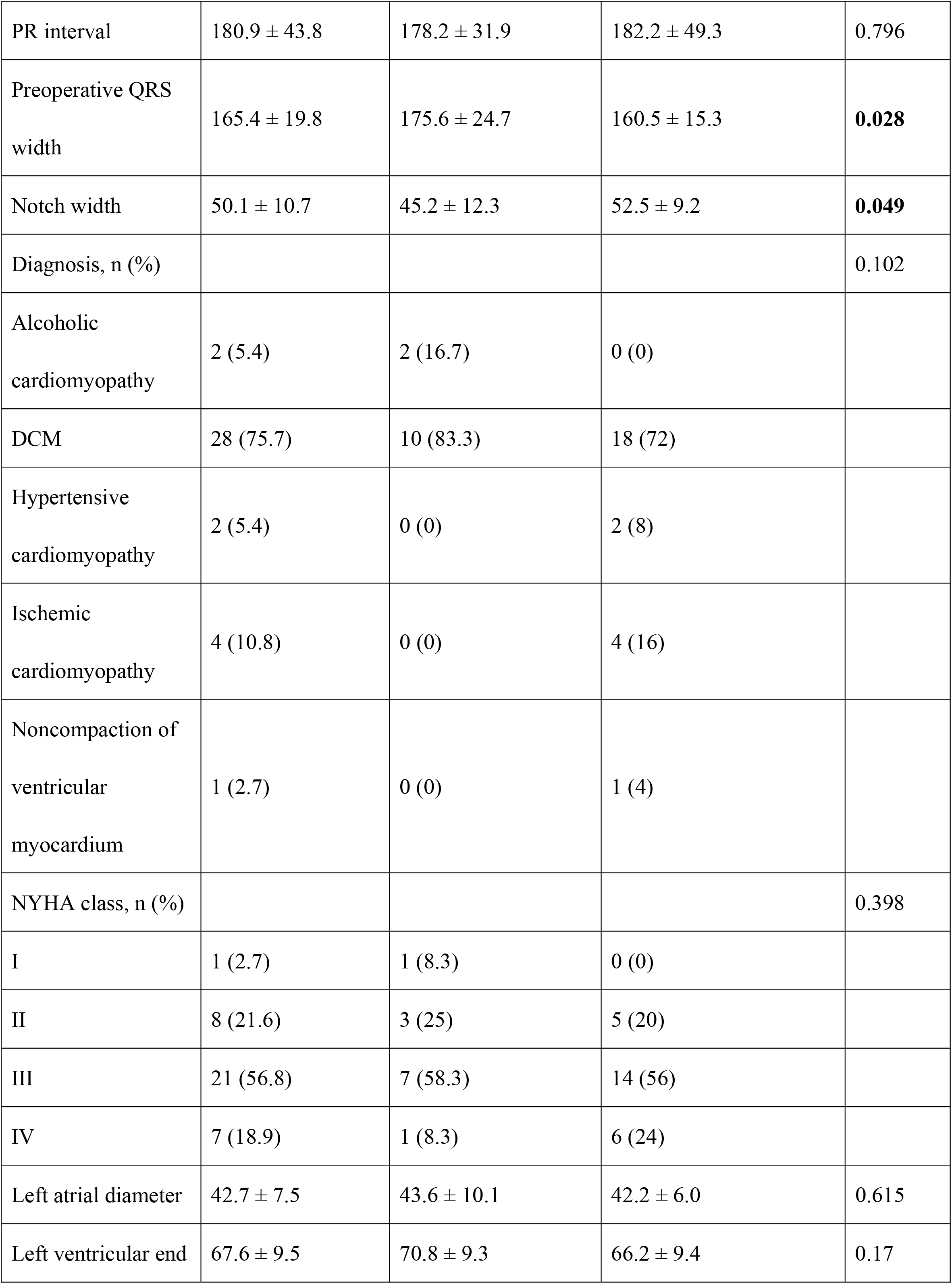
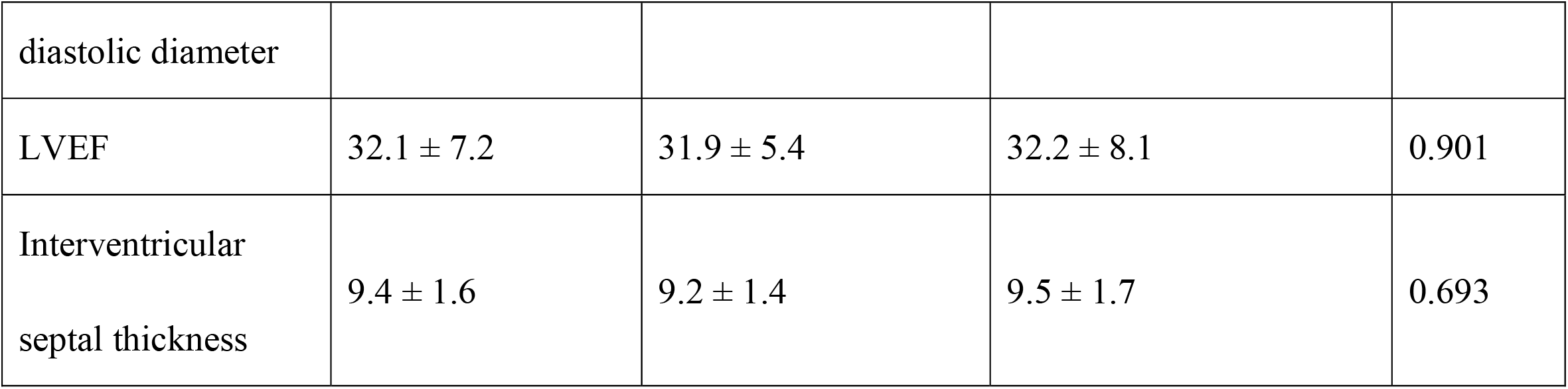
Baseline characteristics of patients analyzed (n = 37).

### Three-dimensional electroanatomical mapping: the mechanism of QRS notch generation differed between the two groups

Left ventricular activation was visualized by high-density three-dimensional electroanatomical mapping. The corresponding relationship between the order of left ventricular activation and the real-time ECG was analyzed. In the t-LBBB group, the start of the QRS wave on the ECG, to the first peak of the notch, corresponded to ventricular electrical activation spreading from the right ventricle to the interventricular septum. The troughs of the notch corresponded to ventricular electrical activation across the left ventricular chamber. The second peak of the notch corresponded to the extension of ventricular electrical activation to the left ventricular free wall. In the f-LBBB group, the left bundle branch conduction was basically normal. When the first peak of the QRS notch appeared, the expansion of ventricular electrical excitation had completed the depolarization of most areas of the right ventricle, septum, and left ventricle—the same as in normal heart depolarization. However, a local slow conduction area existed in the left ventricle, and the depolarization of this part occurred later than in the rest of the myocardium. In addition, the peak was staggered to form a second peak of the notch (**Figure 1, Video 1, Video 2**).

**Figure 1.**
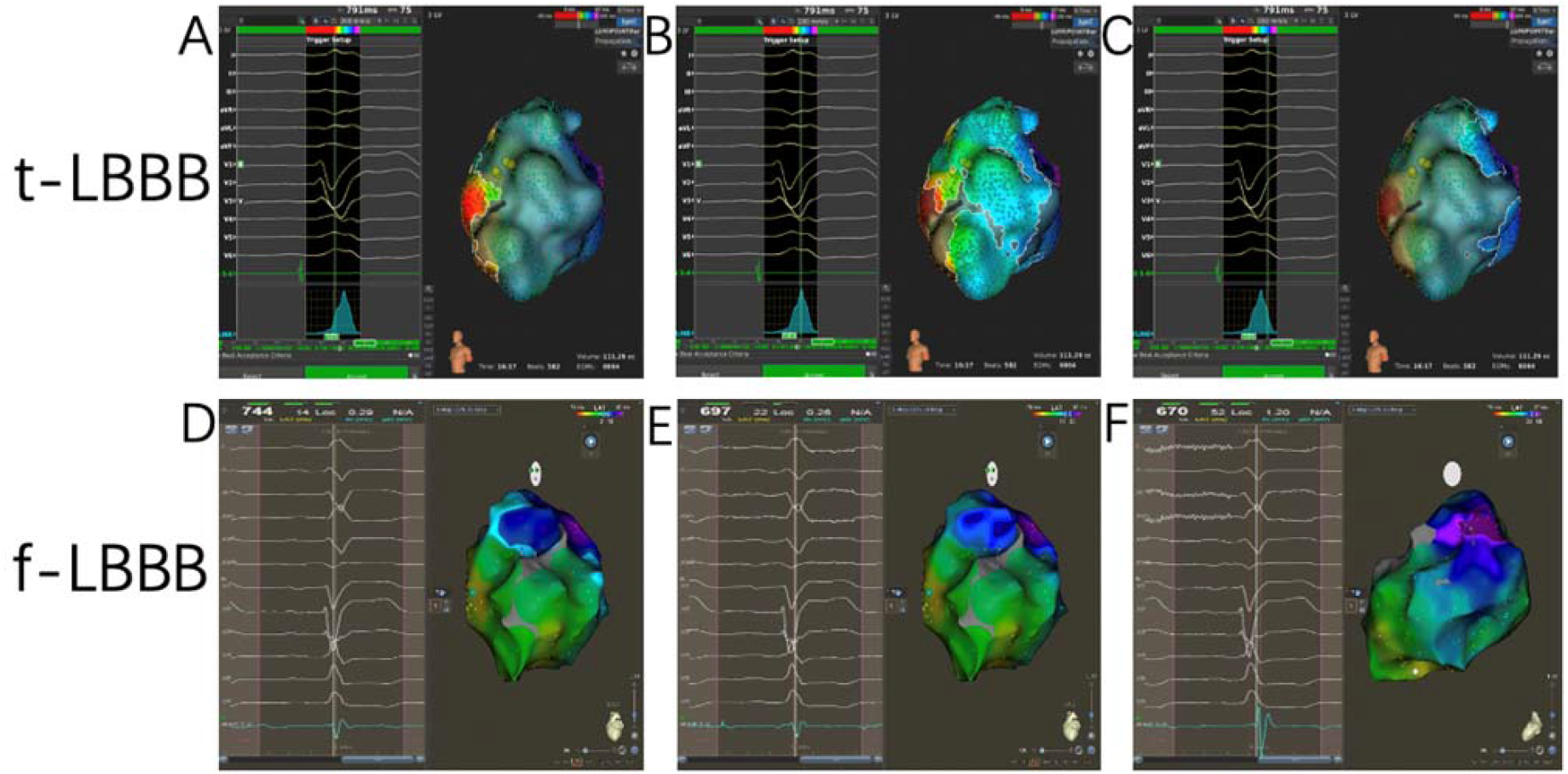
Three-dimensional electroanatomical mapping of the LV. A, B, and C show that electrical excitation reaches the first peak, trough, and second peak of the notch width in t-LBBB. D, E, and F show that electrical excitation reaches the first peak, trough, and second peak of the notch width in f-LBBB.

### The difference in notch width between the t-LBBB group and the f-LBBB group

The QRS notch width of the preoperative surface ECG in the t-LBBB group was longer than that of the f-LBBB group (52.5 ± 9.2 ms in the t-LBBB group vs. 45.2 ± 12.3 ms in the f-LBBB group, *P*<0.05; **Figure 2**).

**Figure 2.**
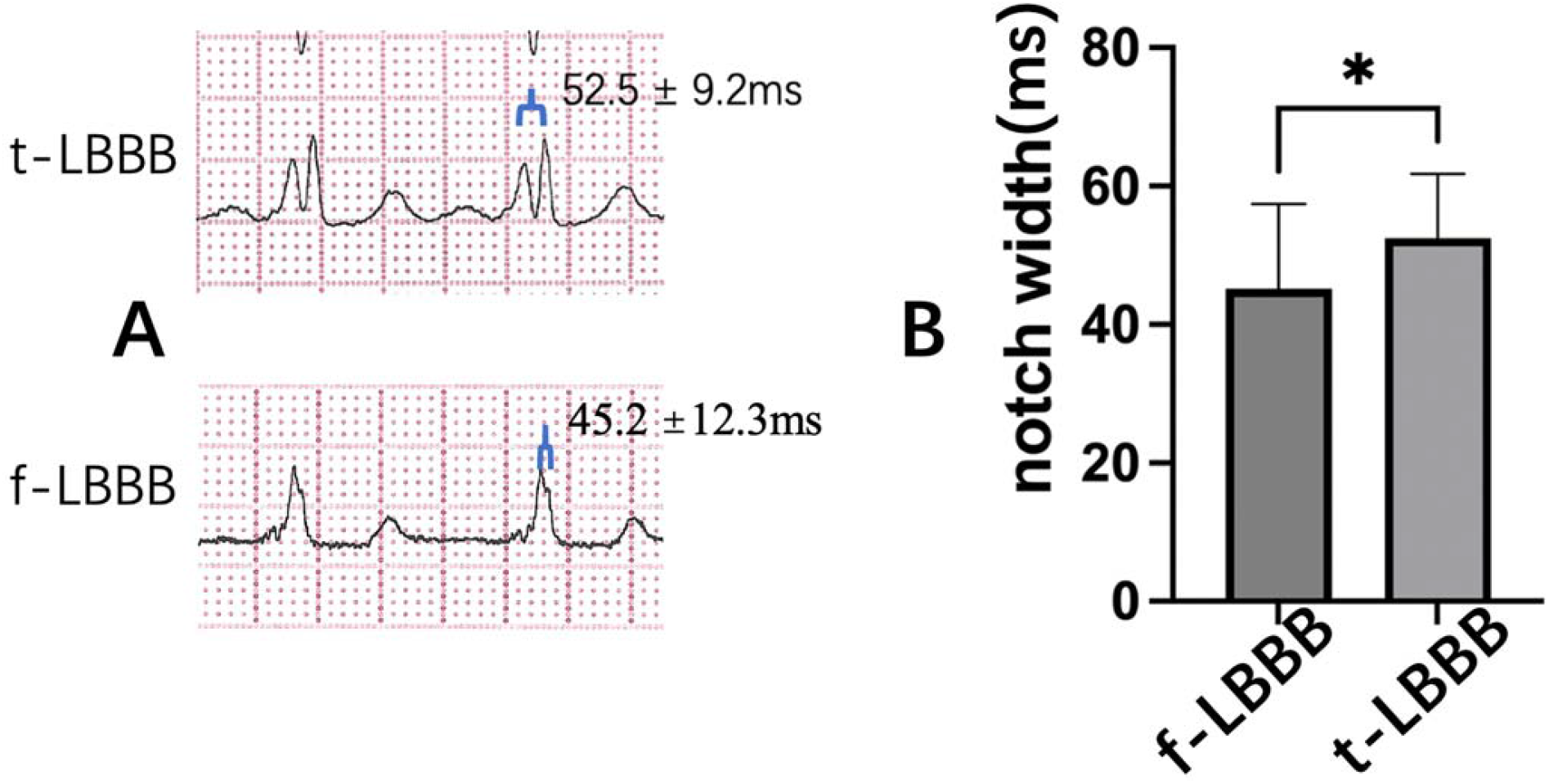
**A**: QRS notch width of t-LBBB and f-LBBB. **B**: t-LBBB group had a wider QRS notch than the f-LBBB group. \**P*<0.05.

No significant difference existed in the left ventricular end-diastolic diameter between the two groups (70.8 ± 9.3 mm in the f-LBBB group vs. 66.2 ± 9.4 mm in the t-LBBB group, *P*>0.05). Considering that the size of the cardiac chamber is related to the QRS duration, we compared the ratio of notch width to left ventricular end-diastolic diameter (Nw/Lvd) and found that Nw/Lvd was significantly greater in the t-LBBB group than in the f-LBBB group (0.81±0.17 ms/mm vs. 0.65±0.19 ms/mm, *P*=0.016; **Figure 3**).

**Figure 3.**
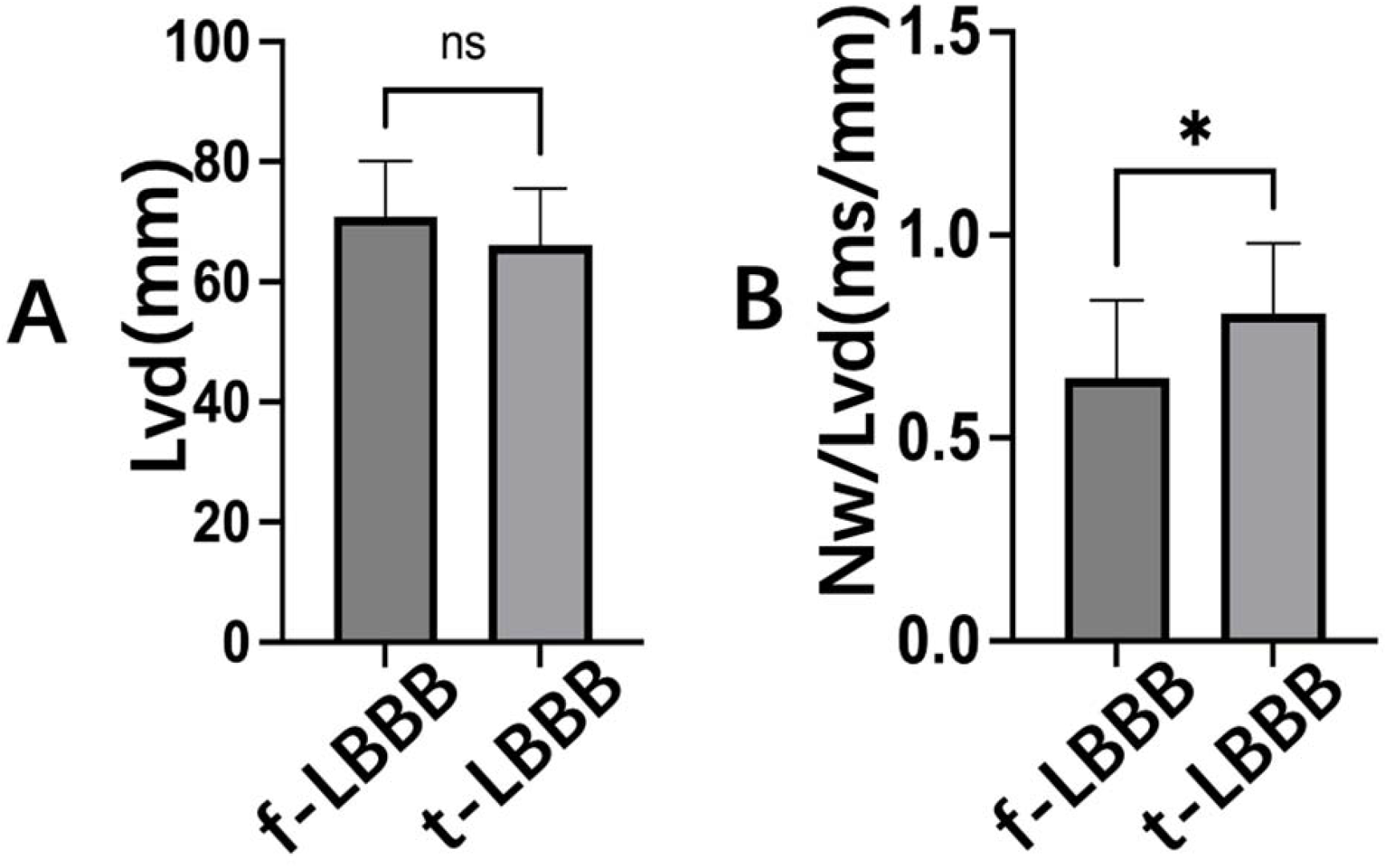
**A**: There was no significant difference in Lvd between the two groups. **B**: The Nw/Lvd in the t-LBBB group was larger than in the f-LBBB group. * *P*<0.05; ns: no significant difference; Nw/Lvd: notch width/left ventricular end-diastole diameter; Lvd: left ventricular end-diastolic diameter.

### Diagnostic value of Nw/Lvd for ROC

We adjusted for confounding factors by logistic regression and found that Nw/Lvd was not affected by age, sex, left ventricular end-diastolic diameter, or history of infarction. It was therefore an independent predictor of t-LBBB (*P*=0.011, OR=3.808, 95% CI 1.36–10.70). The preoperative QRS width was consistent with previous studies and served as an independent predictor (**Table 2**).

**Table 2:**
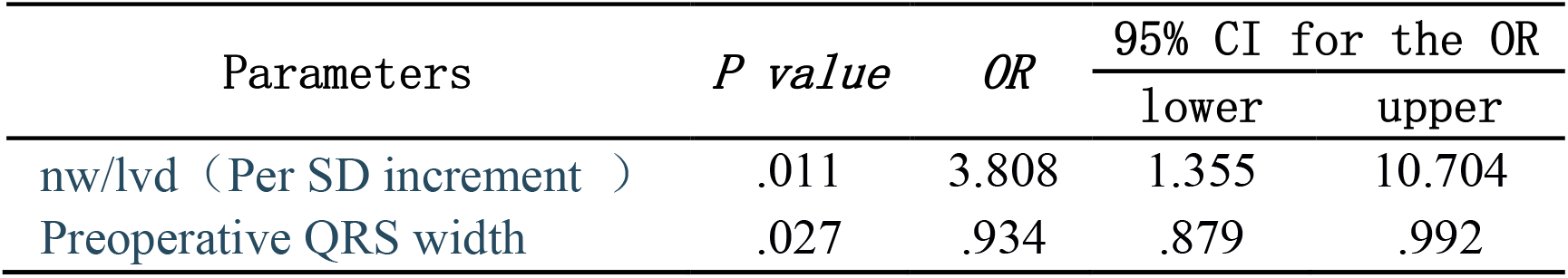
Multivariate logistic regression analysis: predictive value of Nw/Lvd for LBBAP.

ROC analysis showed that the Nw/Lvd ratio was better at diagnosing t-LBBB than Nw alone, informing diagnostic choice. The diagnostic accuracy of t-LBBB by Nw was relatively poor (88% sensitivity, 42% specificity, AUC 0.718, *P*<0.05), and Nw/Lvd demonstrated high sensitivity and specificity (88% sensitivity, 58% specificity, AUC 0.75, *P*<0.05). We obtained an Nw/Lvd cut-off value of 0.56 (**Figure 4**).

**Figure 4.**
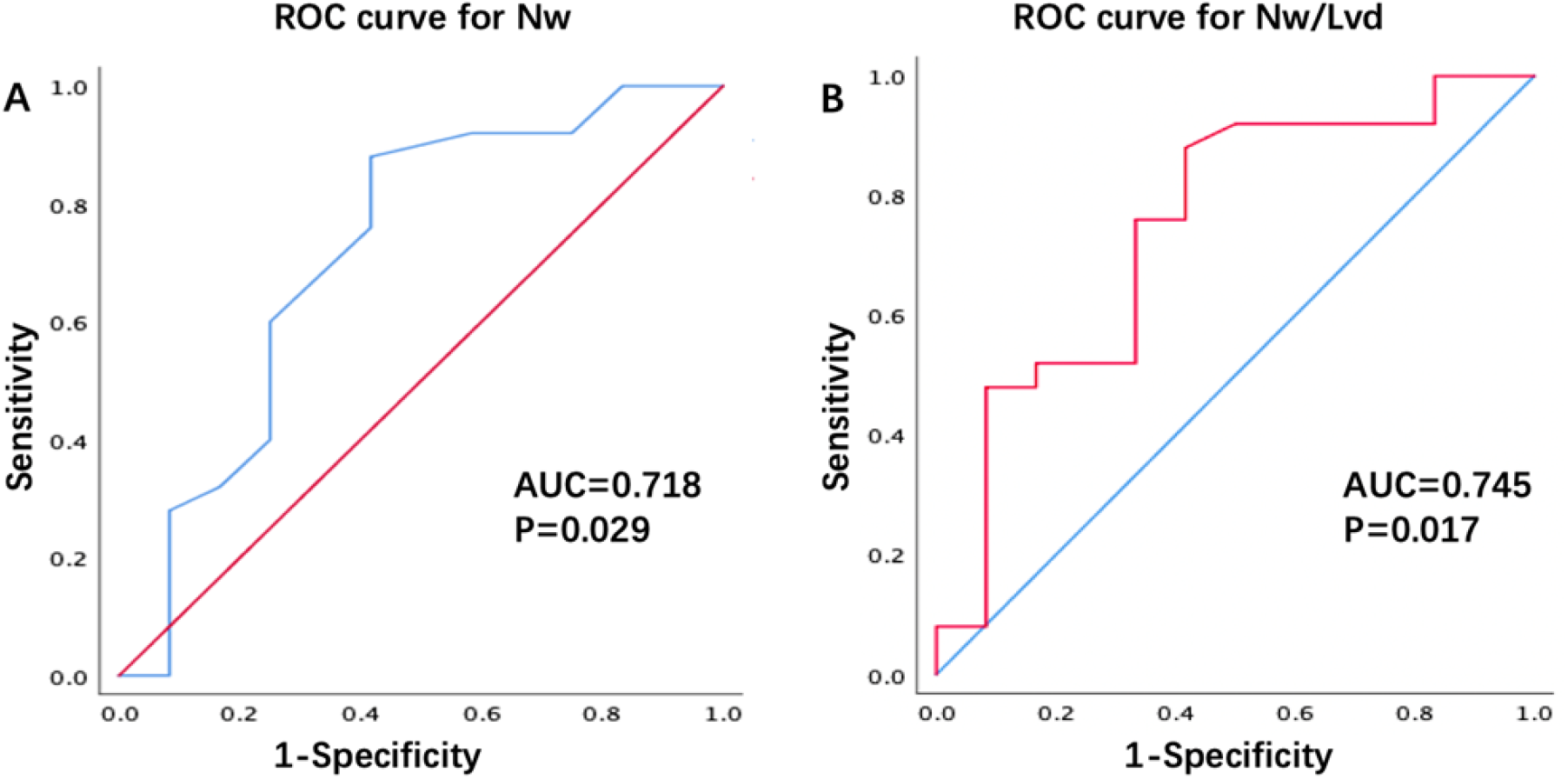
**A**: Receiver operating characteristic curve for Nw in diagnosing response (AUC=0.718, *P*<0.05). **B**: Receiver operating characteristic curve for Nw/Lvd in diagnosing response (AUC=0.745, *P*<0.05). Cut-off value is 0.56. Nw/Lvd: notch width/left ventricular end-diastole diameter; Nw: notch width; AUC: area under the curve.

## Discussion

In this study, we performed intracardiac, high-density three-dimensional electroanatomical mapping in patients who were scheduled to undergo CRT and whose preoperative ECG met the ACC/AHA/HRS guidelines for complete LBBB. We used the mapping results to determine whether an LBBB was present and further subdivided patients into the t-LBBB and f-LBBB groups. Comparing the two groups, we found that (1) the QRS complex notch generation mechanism differed between the two groups, (2) the width of the notch was significantly different, and (3) Nw/LVd could further improve the diagnostic specificity of t-LBBB.

Gaurav et al. performed electroanatomical mapping of the left ventricular septum in patients whose surface ECG showed an LBBB-like ECG (according to the ACC/AHA/HRS guidelines for LBBB diagnostic criteria). They found that only 64% of patients did not record a complete left bundle branch potential conduction. The earliest local ventricular activation point was located in the middle of the septum or the base, rather than the apex (physiologic pattern), which is a t-LBBB (9). However, that study did not further describe the corresponding relationship between the left ventricular activation sequence and real-time ECG in LBBB patients. On the other hand, the current, internationally recognized LBBB diagnostic criteria emphasize that the QRS notch of the lateral wall leads (I, avL, V5, v6) plays an important role in the diagnosis of LBBB (5, 10). Some scholars believe that the notch is formed by the transmission of electrical excitation to the left ventricle and its

U-shaped spread in the left ventricle (11, 12). However, during the process of myocardial depolarization, the fragmented QRS complex reflected by the interruption of conduction continuity or conduction delay of the local ventricular muscle is similar to the notch of LBBB (13). This will affect the judgment of whether to diagnose t-LBBB. Therefore, we speculate that further analysis of this notch width may improve the diagnostic efficiency of t-LBBB.

Intracardiac electroanatomical mapping is used as the gold standard for determining the presence or absence of LBBB and the location of the block (14), which helps to improve the selection of CRT patients (6, 15). In 2004, Auricchio et al. used traditional three-dimensional mapping to describe cardiac chamber activation in patients with LBBB. They found a functional block line in the cardiac chamber, leading to a U-shaped spread of left ventricular electrical excitation and resulting in the formation of notches (16). However, traditional three-dimensional mapping is non-contact balloon mapping, and accuracy is reduced at points that lie 4 cm beyond the balloon. The unipolar recorded electrogram is greatly disturbed, and the identification of potentials in low-voltage areas is limited. In addition, the density of the traditional mapping is not high, and anatomical distortions are prone to occur in complex left ventricular structures. With the emergence of high-density mapping, PentaRay multi-electrode catheters or Orion basket electrodes can be used to map complex left ventricular structures more accurately and precisely. At the same time, it helps us to better understand the ventricular matrix and judge the health of the myocardium (17–19). During our mapping process, we found no evidence of functional block lines. Other studies have also shown that the functional block line of LBBB patients is related to the breakthrough point of the left ventricular endocardium, and its length and shape are very different (16). Therefore, we consider that the appearance of a functional block line may be related to the limitations of traditional three-dimensional mapping.

Through high-density three-dimensional electroanatomical mapping in the cardiac chamber, we analyzed real-time correspondence between the distribution of ventricular electrical excitation and the ECG. We found that, although both were LBBB-like ECGs with notches, the mechanisms of notch formation in t-LBBB and f-LBBB were different. In the t-LBBB group, the trough of the QRS notch on the ECG corresponded to the ventricular electrical excitation across the left ventricular chamber. It is precisely because of the presence of cardiac chambers that the density of the myocardium is small, and the resulting depolarization vector is bound to be small, thus forming troughs. In the f-LBBB group, because a local slow conduction area exists in the left ventricle (probably caused by a myocardial scar), this part is depolarized later than the rest of the myocardium (20, 21) and the second peak of the notch is formed by a staggering peak.

The QRS wave notch is a strong predictor of t-LBBB according to existing guidelines and Strauss criteria (QRS width ≥130 ms in women and ≥140 ms in men; QS or rS in leads V1 and V2; and mid-QRS notching or slurring in two of leads V1, V2, V5, V6, I, and aVL) (5). However, even the most stringent Strauss criteria display a weakness in the diagnosis of LBBB: poor specificity (6). At the same time, when judging complete LBBB, residual LBBB conduction, and intraventricular differential conduction, reliable ECG criteria are lacking (22). Consequently, around one third of patients diagnosed with LBBB by surface ECG may be false positive (7). Our study found that the notch width of the t-LBBB group was significantly larger than that of the f-LBBB group, suggesting that the width of the notch can be used as a new diagnostic criterion for t-LBBB.

Although intracardiac electroanatomical mapping can identify LBBB, it is always an invasive test. However, traditional ECG criteria for diagnosing LBBB have limitations. This study found that the mechanisms of QRS complex notch formation in t-LBBB and f-LBBB are completely different and that the notch width is also significantly different. Therefore, we propose that adding the ratio of notch width to cardiac chamber size (Nw/LVd) to the current diagnostic criteria can further help improve the performance of t-LBBB diagnostic methods. In addition, our previous study confirmed that, in patients with a complete LBBB-like ECG who were scheduled to undergo CRT, intracardiac mapping was performed. Left bundle branch area pacing was performed in the t-LBBB group, and lateral cardiac vein pacing was performed in the f-LBBB group. The postoperative follow-up mapping group had better CRT responsiveness than the non-mapping group (8).

### Limitations

This was a single-center, retrospective study with a small sample size and a limited level of evidence. In addition, this study included only patients presenting with HF who were planning to undergo CRT. Therefore, heterogeneity may exist in complete LBBB between such patients and those without HF or with normal heart size.

### Innovation

This study is one of the few in the world utilizing high-density electroanatomical mapping of LBBB. Further, it informs that the notch width of the QRS wave could be used to improve current guidelines for the diagnosis of LBBB by making up for deficiencies in accuracy.

## Conclusions

In addition to the current diagnostic criteria for LBBB, the predictive measurement of Nw/Lvd is expected to enhance the diagnostic accuracy of t-LBBB.

## Data Availability

Our research has obtained the standards of the ethics committee of the First Affiliated Hospital of Kunming Medical University. We ensure that our research complies with relevant legal and ethical regulations, all experiments were performed in compliance with the First Affiliated Hospital of Kunming Medical University's policy on animal use and ethics.And we have obtained informed consent from everyone who meets the inclusion criteria, and signed relevant documents. We guarantee the authenticity and availability of each patient's data.

## Acknowledgements

We would like to specifically acknowledge Dr. Bao-Tong Hua, Dr. Jing Wang and Dr. Li-Jin Pu as well as other students, nurses and doctors who recruited and helped collect the samples. We would also like to thank the reviewers whose comments/ suggestions helped improve and clarify this manuscript. In addition, we would like to acknowledge and thank all the patients who took part in the study. Without their help this study would not have been possible.

## Abbreviations List

(t-LBBB): true left bundle branch block
(f-LBBB): false left bundle branch block
(Nw): notch width
(LVd): left ventricular end-diastolic diameter
(Nw/LVd): notch width/left ventricular end-diastolic diameter

## Author contributions

BT Hua, J Wang and LJ Pu are in charge of designing research studies.BT Hua, J Wang, LJ Pu, H Li, C Wang, JH Zou are in charge of conducting experiments and acquiring data. C Wang, JH Zou, XX Shao, X Tian and ZX Li are in charge of analyzing data. J Wang and JH Zou are in charge of writing the manuscript. BT Hua and JH Zou are co-first author authors, they contributed equally to this work. J Wang and LJ Pu are co-corresponding authors.

---

**Video 1.**
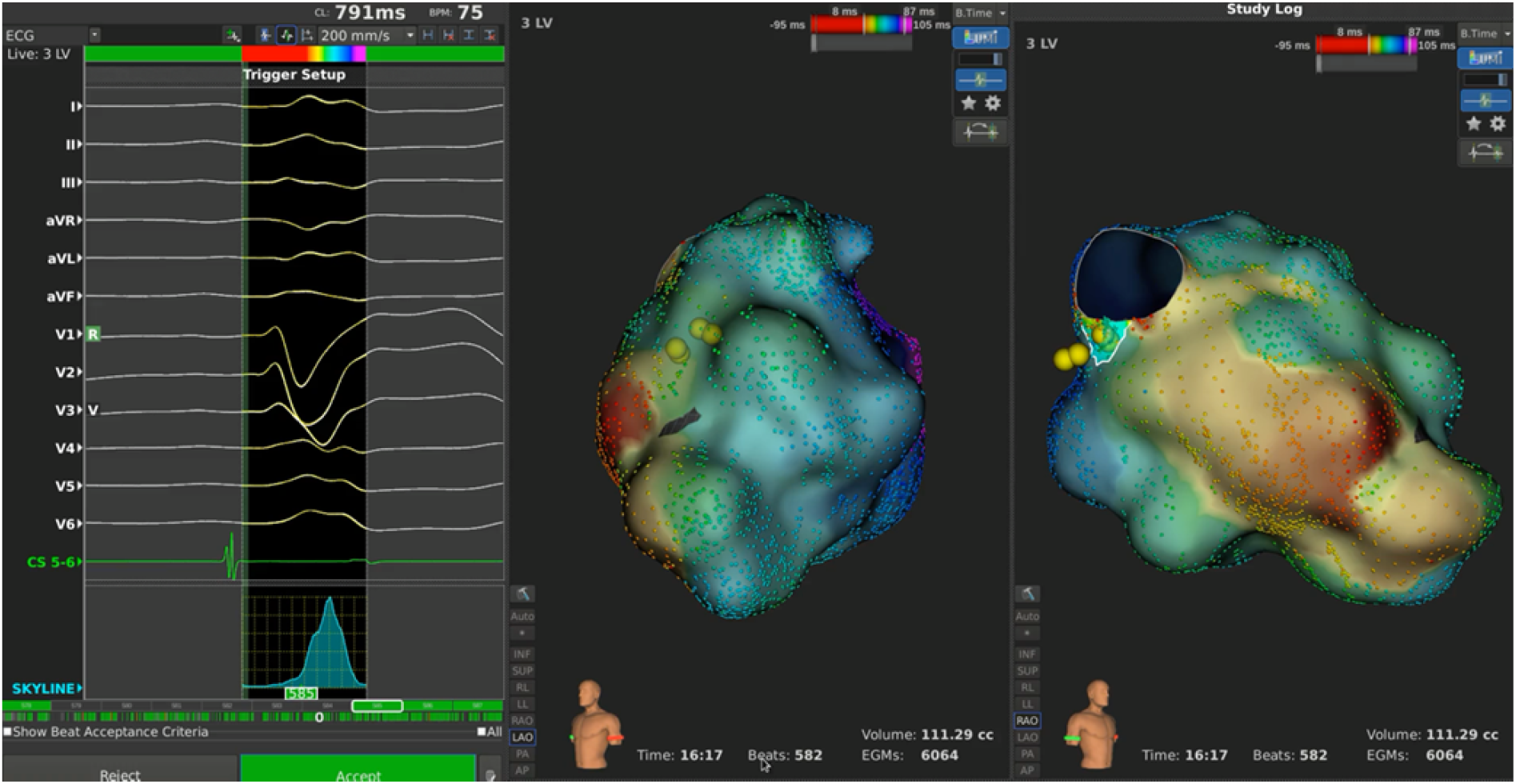
The left ventricular excitation of t-LBBB under high density mapping.

**Video 2.**
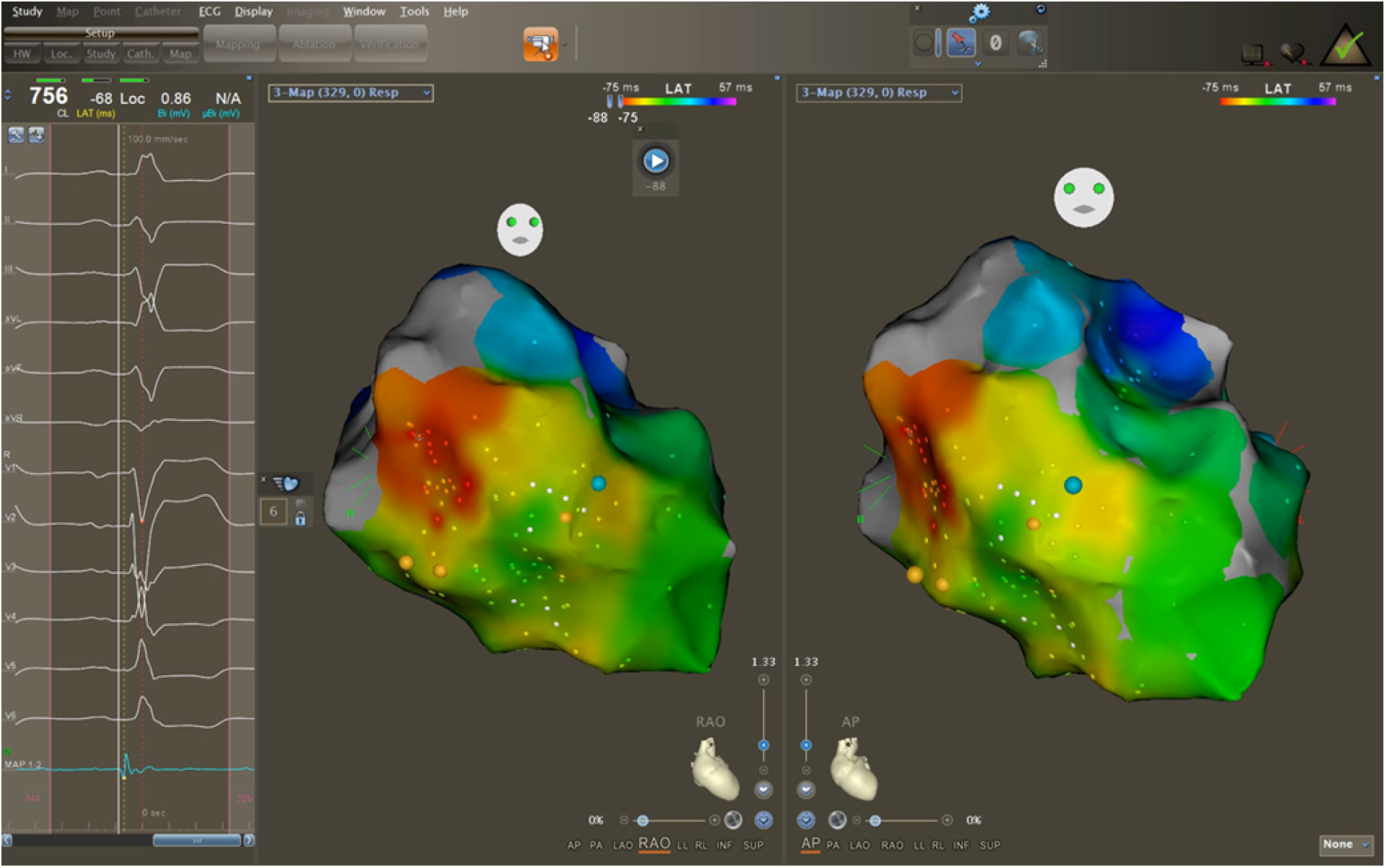
The left ventricular excitation of f-LBBB under high density mapping.

